# REGIONAL DETERMINANTS OF THE EXPANSION OF COVID-19 IN BRAZIL

**DOI:** 10.1101/2020.04.13.20063925

**Authors:** Waldecy Rodrigues, David Nadler Prata, Wainesten Camargo

## Abstract

**Objective:** This study investigates the regional differences in the occurrence of COVID-19 in Brazil and its relationship with climatic and demographic factors by use data from February 26 to April 04, 2020. Methods: A Polynomial Regression Model with cubic adjustments of the number of days of contagion, demographic density, city population and climatic factors was designed and used to explain the spread of COVID-19 in Brazil.

**Main results:** It was evidenced that temperature variation maintains a relationship with the reduction in the number of cases of COVID-19. A variation -3.4% in the number of COVID-19 cases was found for each increase of 1 ° C.

**Conclusion:** There are evidences that the temperature, has a relative effect in the variation in the number of COVID-19’s researched cases. For the reason, it recommends this relationship deserves to be investigated in other tests with more extended time series, wide and with especially non-linear data adjustments.

## INTRODUCTION

COVID-19 is an infectious respiratory disease caused by the SARS-CoV-2 virus, which can cause, depending on general health conditions and anamnesis, the rapid death of those affected by the disease.

A relevant question around the expansion of SARS-CoV-2 is regarding the main climatic or demographic characteristics that allow a larger expansion of this virus. Bukhari and Jameel^1^ report that SARS-CoV-2 is spread rapidly in several countries and has been declared a worldwide pandemic crisis by the World Health Organization. The authors also pointed out that a variant similar to SARS-CoV-2, the influenza virus, was affected by the climate. Today it is unknown whether COVID-19 follows the same scalability (LOWEN^2^; BARRECA & SHIMSHACK^3^).

Bukhari and Jameel^1^ found some important findings in their data, for example, they made an analysis of the climatic patterns of the regions affected by SARS-CoV-2 worldwide until March 22, 2020, with 83% of the tests being performed in non-tropical countries (30N and above) and 90% of SARS-CoV-2 cases were recorded in the same countries within a temperature range of 3 °C to 17 ° C.

However, what caught the attention of these researchers is that in several countries between 30N and 30S, such as Australia, United Arab Emirates, Qatar, Singapore, Bahrain and Taiwan, they performed extensive tests per capita, and the number of positive SARS-CoV-2 per capita is lower in these countries compared to several European countries and the USA.

Therefore, although currently available data is distorted by minimal per capita testing in many tropical countries, it is possible that the climate plays a role in the spread of SARS-CoV-2, which would warrant further investigation. However, with the analysis of the database used in this research, the results were not able to demonstrate with statistical significance the influence of climatic variation in the spread of SARS-CoV-2, and that it would not spread in humid and hot regions with the same speed.

From a study by Wang et al.□ 429 municipalities affected by SARS-CoV-2 around the world, from January 20 to February 4, 2020, where the daily averages, of January 2020, of the temperature (average, minimum and maximum) were calculated. Then, the restricted cubic spline function and the generalized linear mixture model were used to analyse the relationships. The main results of this study pointed to a total of 24,232 confirmed cases in China and 26 countries abroad.

In total, 16,480 cases (68.01%) were from the province of Hubei in China. It was found that in the single-factor model of the highest temperature group, each 1°C increase in the minimum temperature led to a 0.86% reduction in the cumulative number of cases. Thus, the study concluded that, to some extent, the temperature could impact COVID-19 transmission and there may be a better temperature for viral transmission, which may partly explain why the first explosion occurred in Wuhan. It is suggested that countries and regions with the lowest temperature in the world adopt the most stringent control measures to avoid future reversals (WANG ET AL.□).

Brazil is a vast tropical country, with most of its territory located in the range 30N and 30S. But occurring in the range below 30S, it is possible to assess whether there is intervening of the tropical climate variable, which includes the sub-regions called hot and torrid, on the incidence of SARS-CoV-2 on different territories. Brazil is almost entirely composed by Tropical Zone, with an average annual temperature of 24.3 ° C, and a small band of Subtropical Zone, with an average temperature of 18.8 ° C. Are there regional variations in the occurrence of SARS-CoV-2 in these different climatic zones?

Thus, this article aims to investigate the regional differences in the occurrence of COVID-19 in Brazil, its relationship with climatic and demographic factors, based on data of identified cases of COVID-19 from February 26 to April 04, 2020. Brazil has been divided politically under the same structure of the United States of America, comprising of States and each State has its own capital city. In this study, the COVID-19 data was analyzed in the capitals of all the States of Brazil.

## METHODOLOGY

The trajectory of expansion of SARS-CoV-2 in different parts of the planet is necessary knowledge to understand its dynamics under different climatic, demographic and socio-economic conditions. Knowing the behaviour of this virus will be important for future measures of epidemiological control.

For this, we start from the following hypothesis:

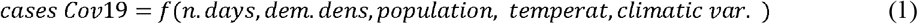

The trend of COVID-19’s cases number are considered as dependent variable (y), and days of contagion (x1), population (x2), demographic density (x3) and temperature (x4) as independent variables. The cubic (y = β0 β1×1 β2×2 β3×3 β4×4) polynomial regression was considered. A significant trend was considered in which the estimated model obtained p value <0.05. For the best model choice, dispersion diagram, coefficient of determination (r2), and residue analysis were also considered. When all the criteria were significant for more than one model and the coefficient of determination was similar, the simplest model was chosen. The analyses were performed using SAS software version 3.8.

The model was generated for testing the climatic variable. The annual average of temperatures was considered, represented in the following function:

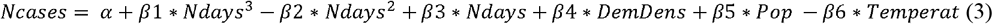

Considering:

*Ncases – number of cases of COVID-19*

*Ndays – number of days since the first case of COVID-19*

*DemDens – demographic density Pop - population*

*Temperat – Annual average temperature in degrees Celsius*

The expected relationship reveals that higher the number of days, thus higher the number of cases of COVID-19, with adjustment in geometric progression, reaching a peak, then a plateau, followed by a subsequent decrease and the possibility of growth recurrences. This adjustment is perfectly compatible with the cubic polynomial function.

It is also expected that higher the concentration of people represented by the demographic density, higher the incidence of COVID-19. In addition, it seeks to test, the Brazilian case, in this initial phase of contamination by SARS-CoV-2, whether the temperature variation is intervening in the volume of occurrences.

In order to operationalize the polynomial regression model, the following steps were taken:

1. Select the explanatory variables, considering the best theorical or empirical relationships;
2. Code the variables;
3. Make scatter plots with all variables, sequentially each pair;
4. Perform the univariate analysis of the independent variables, with their respective analysis of residuals.
5. Develop the correlation matrix to assess the collinearity of the independent variables and define the order of entry of them in the multiple model.
6. Perform the multiple analysis, evaluating the significance of the general model, each of the variables and the increment of each one, through the F test and p-value.
7. Decide on the best model and the best adjustment.

## RESULTS AND DISCUSSION

For data analysis, it was analysed daily data on the occurrence of COVID-19 in the 27 (twenty-seven) capitals of the Brazilian States from February 26 to April 04, 2020□, to verify the hypotheses initially raised by the research, that higher temperatures averages can lead to a lower occurrence of COVID-19.

Thus, to verify the growth of SARS-CoV-2 in the different climatic zones, the third degree linear polynomial model was used from the data of the following capitals of the Brazilian States: Porto Velho (RO), Rio Branco (AC), Manaus (AM)), Boa Vista (RR), Belém (PA), Macapá (AP), Palmas (TO), São Luís (MA), Teresina (PI), Fortaleza (CE), Natal (RN), João Pessoa (PB), Recife (PE), Maceió (AL), Aracaju (SE), Salvador (BA), Belo Horizonte (MG), Vitória (ES), Rio de Janeiro (RJ), Campo Grande (MS), Cuiabá (MT), Goiânia (GO), Brasília (DF), São Paulo (SP), Curitiba (PR), Florianópolis (SC) and Porto Alegre (RS). Those cities that, until the date of the present analysis, April 4, 2020, held about 70% of the cases of COVID-19 in Brazil (Table 1).

**Table 1.**
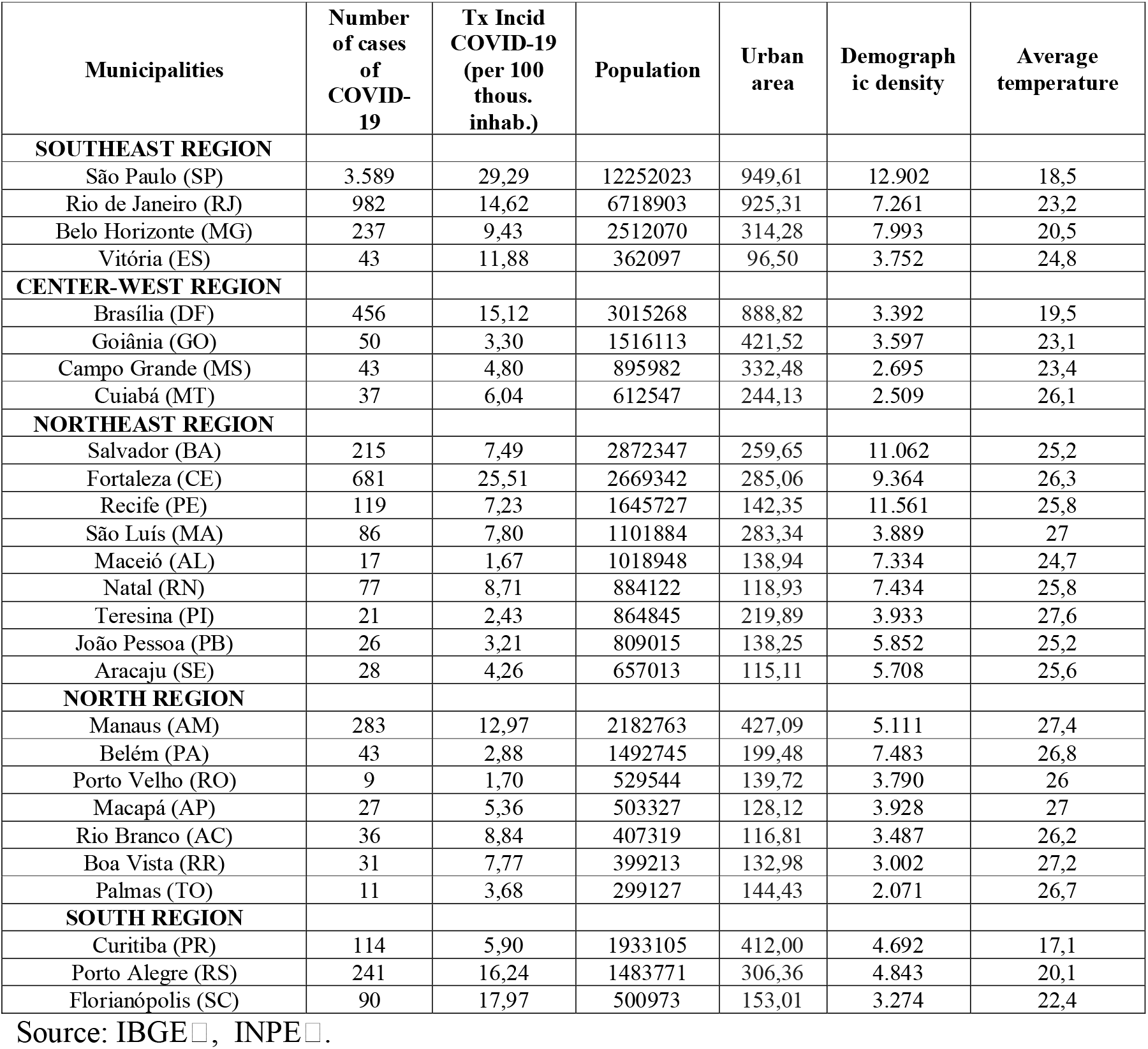
Demographic, climatic and epidemiological information (COVID-19) from the capitals of the Brazilian States - 2020

Correlations were found, on logarithmic scales, between the number of COVID-19 cases in the capitals of the Brazilian States, with the population dimension (0.88) (Graph1), demographic density (0.57) (Graph 2) and with temperature levels (0.55) (Graph 3). All significant results with a *p*-value of less than 1%□.

Once the statistical significance and the correlation between demographic and climatic variables that are involved in the regional differences in the occurrence of COVID-19 were verified, it remains to be seen how the set of demographic and climatic variables act simultaneously in explaining the phenomenon. It starts from the hypothesis that the number of days explains the cases of COVID-19by the number of days with a trajectory in a cubic polynomial function, the demographic density, population and by the temperature of the municipality in question.

The statistical model was generated, as shown below:

| **MODEL (y = log n. case COVID-19)** |  |  |  |  |
| --- | --- | --- | --- | --- |
| **Variables** | **Coefficients** | **Standard error** | **t value** | **p value** |
| Intercept | 0,678220374 | 0,33952182 | 1,98 | 0,0487 |
| Temperat | -0,0479987700 | 0,01277545 | -3,76 | 0,0002 |
| Ndays<sup>3</sup> | 0,0001800549 | 0,00006371 | 2,83 | 0,0049 |
| Ndays<sup>2</sup> | -0,0116853168 | 0,00284904 | -4,10 | < 0,0001 |
| Ndays | 0,3784635405 | 0,03655497 | 10,35 | < 0,0001 |
| DemDens<sup>2</sup> | 0,0000000216 | 0,00000000 | 10,09 | < 0,0001 |
| Population | 0.0000000679 | 0.00000002 | 4,08 | < 0,0001 |
| R<sup>2</sup> | 0,803667 |  |  |  |
| F value | 1814,65 |  |  |  |

The model founds satisfactory “p” and R^2^ values. It means that above 80% of the variations of the dependent variable (Y) are derived from variations of independents variables (X1 …Xn). It is a relatively robust statistical model, with algebraic signs according to the established hypotheses, and, therefore, liable to have its results deepened.

The statistical model plotted on the temporal dynamics of COVID-19 in Brazil, demonstrated a robust dimension of the population and demographic density. The issue of temperature the municipalities also demonstrated that there are strong indications that this variable is also significant, but in smaller dimension.

It was evidenced that the temperature variation maintains a relationship with the reduction in the number of COVID-19 cases. However, this depends on the contagion process in progress. The expected reductions in the number of cases of COVID-19 with the temperature increase in 1° C is -3.4%. (Graph 4)□.

## CONCLUSIONS

In the Brazilian case, the temperature has a reduced magnitude effect despite being intervening in the variation of the number of cases. Other variables have a greater pressure to increase the contagion process - such as the natural process of proliferation of the virus when it enters the cities and the greater number of the population and its densification.

Therefore, cities with higher temperatures may not necessarily present themselves as more protected from the SARS-CoV-2 than those with lower temperatures. The variations found in this work indicate that as the contagion process in cities advances, the reductions become smaller at different average temperatures.

It is noteworthy that the result found in this work, the COVID-19’s cases variation is -3.4% for each increase of 1 ° C. International studies relevant to the relationship between temperature and the occurrence of the SARS-CoV-2 carried out by Bukhari and Jameel^1^, Wang et al. □ and Bogoch□ in colder climatic zones already pointed out that a reduction in temperature may contribute to a more significant proliferation of the virus. However, an increase in temperature is not necessarily a major barrier to its expansion.

## Data Availability

All data used by the survey are open to the public.

https://brasil.io/dataset/covid19/caso?search=&date=2020-04-01&state=TO&city=&place_type=&is_last=&city_ibge_code=&order_for_place=

